## supplementary tables for "Epidemiology of alcohol use and alcohol use disorder among female sex workers in Mbeya City, Tanzania"

**Table 1: Reported alcohol use in the past 12 months among Female Sex Workers and associated factors by bivariate analysis.**

| Characteristic | Categories | Alcohol Use in past 12 months | | | | Total | P-value |
| --- | --- | --- | --- | --- | --- | --- | --- |
|  |  | **No** | | **Yes** | |  |  |
|  |  | n | % | n | % | n |  |
| Residency/location | Mbeya City | 23 | 15.1 | 129 | 84.9 | 152 | 0.234 |
|  | Others | 0 | 0 | 8 | 100 | 8 |  |
| Age (years) | 18-24 years | 7 | 10.3 | 61 | 89.7 | 68 | 0.206 |
|  | 25 and above | 16 | 17.4 | 76 | 82.6 | 92 |  |
| Religion | Muslims | 4 | 10 | 36 | 90 | 40 | 0.770 |
|  | Roman Catholics | 11 | 16.4 | 56 | 83.6 | 67 |  |
|  | Protestants | 7 | 16.3 | 36 | 83.7 | 43 |  |
|  | Others | 1 | 10 | 9 | 90 | 10 |  |
| Level of education | Incomplete primary school and never | 6 | 17.1 | 29 | 82.9 | 35 | 0.218 |
|  | Primary school | 11 | 19.3 | 46 | 80.7 | 57 |  |
|  | Secondary school and above | 6 | 8.8 | 62 | 91.2 | 68 |  |
| Marital status | Single | 11 | 8.9 | 113 | 91.1 | 124 | <0.001 |
|  | In a relationship (including married and cohabiting) | 12 | 33.3 | 24 | 66.7 | 36 |  |
| Income | Median (90000 and below) | 16 | 20.3 | 63 | 79.7 | 79 | 0.036 |
|  | Above median income (>90000) | 7 | 8.6 | 74 | 91.4 | 81 |  |
| Dependents | Yes | 13 | 12.3 | 93 | 87.7 | 106 | 0.286 |
|  | No | 10 | 18.5 | 44 | 81.5 | 54 |  |
| Area of sleeping | Home (Owned or rented) | 7 | 9.7 | 65 | 90.3 | 72 | 0.264 |
|  | Hotel/Street/Brothel | 4 | 15.4 | 22 | 84.6 | 26 |  |
|  | Sleep at Friends and relatives | 12 | 19.7 | 49 | 80.3 | 61 |  |
| Drinking alcohol before sex | Yes | 4 | 5.9 | 64 | 94.1 | 68 | 0.008 |
|  | No | 19 | 20.7 | 73 | 79.3 | 92 |  |
| Condom Use | Yes | 6 | 18.2 | 27 | 81.8 | 33 | 0.484 |
|  | No | 17 | 13.4 | 110 | 86.6 | 127 |  |
| STI | Yes | 2 | 7.1 | 26 | 92.9 | 28 | 0.230 |
|  | No | 21 | 15.9 | 111 | 84.1 | 132 |  |
| Testing for HIV | Yes | 21 | 15.2 | 117 | 84.8 | 138 | 0.447 |
|  | No | 2 | 9.1 | 20 | 90.9 | 22 |  |
| Sexual Partners in past 12 months | 1-10 partners | 15 | 18.8 | 65 | 81.3 | 80 | 0.115 |
|  | More than 10 partners | 8 | 10 | 72 | 90 | 80 |  |
| Sexual Partner in past 30 days | None | 2 | 33.3 | 4 | 66.7 | 6 | 0.393 |
|  | One | 1 | 11.1 | 8 | 88.9 | 9 |  |
|  | More than one | 20 | 13.8 | 125 | 86.2 | 145 |  |
| Regretful sex after alcohol use (N=149) | Yes | 5 | 5.7 | 82 | 94.3 | 87 | <0.001 |
|  | No | 18 | 29 | 44 | 71 | 62 |  |

**Table 2: Reported alcohol use in the past 30 days among Female Sex Workers and associated factors by bivariate analysis**

| Characteristic | Categories (N=160) | Past 30 days alcohol use | | | | Total | P value |
| --- | --- | --- | --- | --- | --- | --- | --- |
|  |  | **No** | | **Yes** | |  |  |
|  |  | n | % | n | % | n |  |
| Location | Mbeya City | 24 | 15.8 | 128 | 84.2 | 152 | 0.223 |
|  | Others | 0 | 0 | 8 | 100 | 8 |  |
| Age (years) | 18-24 years | 8 | 11.8 | 60 | 88.2 | 68 | 0.324 |
|  | 25 and above | 16 | 17.4 | 76 | 82.6 | 92 |  |
| Religion | Muslims | 6 | 15 | 34 | 85 | 40 | 0.523 |
|  | Roman Catholics | 12 | 17.9 | 55 | 82.1 | 67 |  |
|  | Protestants | 6 | 14 | 37 | 86 | 43 |  |
|  | Others | 0 | 0 | 10 | 100 | 10 |  |
| Level of Education | Incomplete primary school and never | 7 | 20 | 28 | 80 | 35 | 0.170 |
|  | Primary school | 11 | 19.3 | 46 | 80.7 | 57 |  |
|  | Secondary school and above | 6 | 8.8 | 62 | 91.2 | 68 |  |
|  | Total | 24 | 15 | 136 | 85 | 160 |  |
| Marital Status | Single | 12 | 9.7 | 112 | 90.3 | 124 | <0.001 |
|  | In a relationship (including married and cohabiting) | 12 | 33.3 | 24 | 66.7 | 36 |  |
| Income | Median (90000 and below) | 18 | 22.8 | 61 | 77.2 | 79 | 0.006 |
|  | Above median income (>90000) | 6 | 7.4 | 75 | 92.6 | 81 |  |
| Dependents | Yes | 12 | 11.3 | 94 | 88.7 | 106 | 0.068 |
|  | No | 12 | 22.2 | 42 | 77.8 | 54 |  |
| Place of sleeping (N=159) | Home (Owned or rented) | 8 | 11.1 | 64 | 88.9 | 72 | 0.225 |
|  | Hotel/Street/Brothel | 3 | 11.5 | 23 | 88.5 | 26 |  |
|  | Sleep at Friends and relatives | 13 | 21.3 | 48 | 78.7 | 61 |  |
| Drinking alcohol before sex | Yes | 5 | 7.4 | 63 | 92.6 | 68 | 0.020 |
|  | No | 19 | 20.7 | 73 | 79.3 | 92 |  |
| Condom Use | Yes | 6 | 18.2 | 27 | 81.8 | 33 | 0.566 |
|  | No | 18 | 14.2 | 109 | 85.8 | 127 |  |
| STI | Yes | 1 | 3.6 | 27 | 96.4 | 28 | 0.062 |
|  | No | 23 | 17.4 | 109 | 82.6 | 132 |  |
| HIV testing | Yes | 23 | 16.7 | 115 | 83.3 | 138 | 0.139 |
|  | No | 1 | 4.5 | 21 | 95.5 | 22 |  |
| Sexual Partners in past 12 months | 1-10 partners | 16 | 20 | 64 | 80 | 80 | 0.077 |
|  | More than 10 partners | 8 | 10 | 72 | 90 | 80 |  |
| Sexual Partner in past 30 days | None | 3 | 50 | 3 | 50 | 6 | 0.049 |
|  | One | 1 | 11.1 | 8 | 88.9 | 9 |  |
|  | More than one | 20 | 13.8 | 125 | 86.2 | 145 |  |
| Regretful sex after alcohol use (N=149) | Yes | 3 | 3.4 | 84 | 96.6 | 87 | <0.001 |
|  | No | 20 | 32.3 | 42 | 67.7 | 62 |  |

**Table 3: Reported High Episodic** Drinking among **Female Sex Workers in and associated factors by bivariate analysis.**

| Characteristic | Categories (N=137) | High Episodic Drinking | | | | Total | P value |
| --- | --- | --- | --- | --- | --- | --- | --- |
|  |  | **No** | | **Yes** | |  |  |
|  |  | n | % | n | % | n |  |
| Location | Mbeya City | 29 | 22.5 | 100 | 77.5 | 129 | 0.011 |
|  | Others | 5 | 62.5 | 3 | 37.5 | 8 |  |
| Age (years) | 18-24 years | 18 | 29.5 | 43 | 70.5 | 61 | 0.255 |
|  | 25 and above | 16 | 21.1 | 60 | 78.9 | 76 |  |
|  | Total | 34 | 24.8 | 103 | 75.2 | 137 |  |
| Religion | Muslims | 11 | 30.6 | 25 | 69.4 | 36 | 0.561 |
|  | Roman Catholics | 12 | 21.4 | 44 | 78.6 | 56 |  |
|  | Protestants | 10 | 27.8 | 26 | 72.2 | 36 |  |
|  | Others | 1 | 11.1 | 8 | 88.9 | 9 |  |
| Level of education | Incomplete primary school and never | 10 | 34.5 | 19 | 65.5 | 29 | 0.004 |
|  | Primary school | 17 | 37 | 29 | 63 | 46 |  |
|  | Secondary school and above | 7 | 11.3 | 55 | 88.7 | 62 |  |
| Marital Status | Single | 22 | 19.5 | 91 | 80.5 | 113 | 0.002 |
|  | In a relationship (including married and cohabiting) | 12 | 50 | 12 | 50 | 24 |  |
| Income | Median (90000 and below) | 25 | 39.7 | 38 | 60.3 | 63 | <0.001 |
|  | Above median income | 9 | 12.2 | 65 | 87.8 | 74 |  |
| Dependents | Yes | 13 | 14 | 80 | 86 | 93 | <0.001 |
|  | No | 21 | 47.7 | 23 | 52.3 | 44 |  |
| Place of sleeping  (N=136) | Home (Owned or rented) | 19 | 29.2 | 46 | 70.8 | 65 | 0.54 |
|  | Hotel/Street/Brothel | 5 | 22.7 | 17 | 77.3 | 22 |  |
|  | Sleep at Friends and relatives | 10 | 20.4 | 39 | 79.6 | 49 |  |
| Sex after alcohol use | Yes | 15 | 23.4 | 49 | 76.6 | 64 | 0.726 |
|  | No | 19 | 26 | 54 | 74 | 73 |  |
| Condom Use | Yes | 13 | 48.1 | 14 | 51.9 | 27 | 0.002 |
|  | No | 21 | 19.1 | 89 | 80.9 | 110 |  |
| STI | Yes | 9 | 34.6 | 17 | 65.4 | 26 | 0.199 |
|  | No | 25 | 22.5 | 86 | 77.5 | 111 |  |
| HIV testing | Yes | 21 | 17.9 | 96 | 82.1 | 117 | <0.001 |
|  | No | 13 | 65 | 7 | 35 | 20 |  |
| Number of sexual partners in past 12 months | 1-10 partners | 29 | 44.6 | 36 | 55.4 | 65 | <0.001 |
|  | More than 10 partners | 5 | 6.9 | 67 | 93.1 | 72 |  |
| Number of sexual partners in past 30 days | None | 3 | 75 | 1 | 25 | 4 | 0.039 |
|  | One | 3 | 37.5 | 5 | 62.5 | 8 |  |
|  | More than one | 28 | 22.4 | 97 | 77.6 | 125 |  |
| Regretful sex after alcohol use(N=126) | Yes | 13 | 15.9 | 69 | 84.1 | 82 | 0.070 |
|  | No | 13 | 29.5 | 31 | 70.5 | 44 |  |
